# Correction of Artifacts Induced by B_0_ Inhomogeneities in Breast MRI using Reduced Field-of-View Echo-Planar Imaging and Enhanced Reverse Polarity Gradient Method

**DOI:** 10.1101/2020.03.31.20048900

**Authors:** Ana E. Rodríguez-Soto, Helen Park, Dominic Holland, Kathryn E. Keenan, Hauke Bartsch, Joshua Kuperman, Anne M. Wallace, Michael Hahn, Haydee Ojeda-Fournier, Anders M. Dale, Rebecca Rakow-Penner

**Author notes:** **Corresponding Author:** Rebecca Rakow-Penner, MD, PhD, Department of Radiology, University of California, San Diego, 9400 Campus Point Drive #7316, La Jolla, CA 92093. NIST required disclaimer: Certain commercial instruments and software are identified to specify the experimental study adequately. This does not imply endorsement by NIST or that the instruments and software are the best available for the purpose.

## Abstract

**Purpose:** Diffusion weighted (DW) echo-planar imaging (EPI) is prone to geometric and intensity distortions due to B_0_ inhomogeneities. Pulse sequences that excite spins within a reduced field-of-view (FOV) in the phase encoding (PE) direction have been developed to decrease such distortions. In addition, use of the reverse polarity gradient (RPG) method, a retrospective approach to correct distortion artifacts, has been shown to improve the localization of tumor lesions. The purpose of this work was to evaluate the performance of reduced-FOV acquisition and RPG in decreasing distortion artifacts for breast imaging.

**Methods:** EPI data were acquired with full and reduced-FOV in a breast phantom and in a group of 170 women at 3T. The performance of RPG in correcting distortion artifacts in EPI data was evaluated using the mutual information (MI) metric between EPI and anatomical low-distortion images before and after distortion correction.

**Results:** RPG corrected distortions by 61% in full-FOV EPI and 48% in reduced-FOV EPI in a breast phantom. In patients, MI increased on average 13±8% and 8±6% for both full and reduced-FOV EPI data after distortion correction, respectively. The 95^th^ percentile and maximum displacement between uncorrected and corrected full-FOV EPI datasets were 0.8±0.3cm and 1.9±0.3cm, and for reduced-FOV were 0.4±0.2cm and 1.3±0.3cm.

**Conclusion:** Minimal distortion was achieved with RPG applied to reduced-FOV EPI data. RPG improved distortions for full-FOV, but with more modest improvements and limited correction near the nipple.

## Introduction

Breast cancer is the most common cancer in American women after skin cancer.^1^ It is estimated that 1 in 8 U.S. women will develop a form of breast cancer in their lifetime, emphasizing the importance of effective and accurate screening practices.^2^ Breast magnetic resonance imaging (MRI) is currently used for screening women with high-risk of breast cancer,^3^ evaluating new breast cancer diagnosis, and other indications such as response to neoadjuvant chemotherapy.^4^ Clinical breast MRI protocols include contrast enhanced (CE)-MRI, in which intravenous contrast agents are administered to visualize vascular patterns, such as tumor angiogenesis. The use of contrast agents is contraindicated in pregnant women. In addition, brain and bone depositions of such contrast agents have been reported in patients with cumulative exposure with unknown sequelae.^5-7^ Hence, there is a need for the development of contrast-free MRI protocols and thus, reduce the number of patients who undergo contrast exposure. Diffusion-weighted (DW)-MRI has been incorporated into breast MRI protocols and demonstrates great potential to become a contrast-free diagnostic tool for breast cancer.^8^ Yet its widespread clinical use has been severely hampered by distortion artifacts that limit the accurate location of lesions.

Diffusion-weighted MRI is commonly acquired using single-shot echo-planar imaging (EPI) readout, which minimizes acquisition time by continuously traversing k-space after a single radio-frequency excitation pulse. In EPI, the bandwidth in the phase encoding (PE) direction is much lower than in the frequency encoding direction. Consequently, spatial and intensity distortions occur in regions of B_0_ inhomogeneities arising at air-tissue interfaces (e.g. anterior chest wall and breast tissue boundaries) due to magnetic susceptibility difference. Because of the low bandwidth in the PE direction, these artifacts predominantly appear in the PE direction.^9^ Geometrically distorted DW-MRI limits the accurate location of tumor lesions. In breast imaging, these artifacts are pronounced due to off-isocenter scanning and complexity of the anatomy.

Efforts to decrease geometric distortions present in DW-EPI for breast imaging include both prospective and retrospective methods.^10-13^ A prospective approach uses pulse sequences to excite spins within a reduced field-of-view (FOV) in the PE direction, resulting in a rectangular FOV. The magnitude of the distortion artifact is reduced by minimizing EPI readout duration; therefore, shortening the time during which spin dephasing evolves due to the contribution of B_0_-inhomogeneities.^14^ The use of reduced-FOV in DW-EPI was shown to increase in-plane resolution and reduce geometric distortions, initially in the spinal cord^14^, and subsequently in other organs such as prostate^15^, pancreas^16^, kidneys^17^ and breast^13^.

Most published retrospective distortion correction algorithms rely on field mapping to estimate local off-resonance across images.^18^ In contrast, Holland et al developed a distortion correction method that exploits the symmetry of the artifacts in the positive and negative PE trajectories: regions with spins that become compressed in images collected with a given PE trajectory polarity (e.g. anterior-posterior, A/P) will be expanded in images collected with the opposite trajectory polarity (e.g. posterior-anterior, P/A). This approach, termed reverse polarity gradient (RPG) requires the acquisition of two non-diffusion weighted, b=0 s/mm^2^ volumes, one with positive and one with negative phase encoding direction.^9^ These data are used to estimate the deformation field that minimizes the difference between both volumes, and to unwarp the diffusion-weighted volumes. The use of RPG improved the localization of brain, prostate, and breast tumor lesions.^19,20^ A version of RPG is currently offered for brain applications only by GE (General Electric Healthcare, Milwaukee, Wisconsin, USA).

Reduced-FOV EPI produces images with lesser geometric distortions, but does not eliminate them; however, there is a continued need for improvement in contrast-free clinical breast diffusion imaging. Given the importance of accurate cancer localization in breast cancer screening and monitoring practices, investigating the application of reduced-FOV EPI and RPG in breast MRI is necessary. Data describing the combined performance of these methods are lacking. Therefore, the purpose of this work is to evaluate the sole and combined performance of these prospective and retrospective distortion correction methods for breast MRI.

## Methods

### Phantom Studies

A breast phantom designed by the University of California San Francisco (UCSF) and the National Institute of Standards and Technology (NIST)^21^ was scanned using a 3T MRI scanner (MR750, GE Healthcare, Milwaukee, Wisconsin, USA) and a breast array coil. Only b=0 s/mm^2^ volumes of DW-MRI acquisitions and T_2_ fast spin echo (FSE) images were used. Pulse sequence parameters: (1) *T*_*2*_ *fat suppressed FSE*: TE/TR=107/4520ms, flip angle of 111°, FOV=320×320mm^2^, acquisition matrix 512×320, reconstruction matrix 512×512, voxel size=0.625×0.625×5mm^3^; (2) *full-FOV DW-MRI*: TE/TR=82/9000ms, FOV=320×320mm^2^, acquisition matrix 96×96, voxel size=2.5×2.5×5mm^3^, spectral attenuated inversion recovery (SPAIR) fat suppression, PE direction left-right (L/R), no parallel imaging; and (3) *reduced-FOV DW-MRI*: TE/TR=82/9000ms, FOV=160×320mm^2^, acquisition matrix 48×96, reconstruction matrix 128×128, voxel size=2.5×2.5×5mm^3^, SPAIR fat suppression, PE direction A/P, no parallel imaging. The pulse sequence parameters between full and reduced-FOV DW-MRI data were identical except for the PE direction and the FOV in the PE direction. Although the expected TE should be shorted in reduced-FOV EPI this is not the case because a longer period is needed to apply the 2D RF excitation pulse.

The breast phantom contains a polycarbonate plate with a grid of circles spaced at 1.5 cm intervals. The spatial discrepancy in the center of each circle between EPI and T_2_ FSE images was measured to quantify the magnitude of the initial distortion and to evaluate the performance of RPG.

### In vivo Studies

In this retrospective study, diagnostic and surveillance MRI was performed in 170 women (52.4±13.4 years old) between January 2016 and July 2019, as part of the standard-of-care MRI protocol at our institution. Patients were scanned in the prone position using identical setup and pulse sequence parameters as described above (except in some cases FOV was 360×360mm^2^ or 180×360mm^2^ with the same acquisition matrix and increased voxel size). Full-FOV DW-MRI were acquired with PE in the L/R direction to minimize cardiac motion artifacts. However, due to the rectangular geometry of the reduced-FOV, these images were acquired with PE in the A/P direction. This retrospective study was approved by the Institutional Review Board at the University of California, San Diego (UCSD). The MRI data in the present study were acquired using the standard of care clinical imaging protocol at UCSD.

### Distortion Correction Algorithm

Both full and reduced-FOV EPI volumes were collected in the positive and negative PE trajectories to retrospectively correct for B_0_-inhomogeneity induced artifacts with RPG. The RPG approach exploits the symmetry of artifacts in the positive and negative PE trajectory acquisitions.^19^ Briefly, the distortion correction algorithm is as follows: 1) positive and negative images are blurred with a Gaussian kernel of determined width (initial kernel size is user-defined), 2) blurred positive and negative images are registered to each other, 3) the displacement field that corrects both images is estimated and used to inform and update the deformation field that will correct original positive and negative volumes based on the following a cost function *f*, and 4) the width of the Gaussian kernel used in step 1 is reduced. Steps 1-4 are repeated until the kernel width is zero. The cost function *f* is defined as:

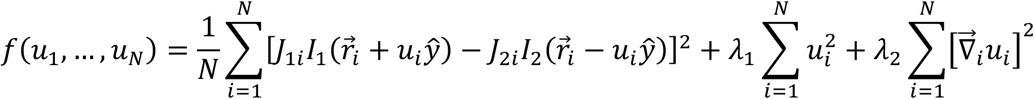

where *u*_*N*_ is the displacement shift of the i^th^ voxel located at 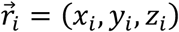 that corrects the distorted images in the positive and negative PE trajectories *I*_1_ and *I*_2_, respectively. While *J*_1_ and *J*_2_ are the Jacobian of the transformation between uncorrected and distortion corrected images, and λ_1_ and λ_2_ are regularization parameters of the model that control the variability of the three-dimensional displacement field.

Each EPI dataset was distortion corrected with RPG using all the combinations of Gaussian kernel width (60, 70 and 80 pixels) and λ_2_ (500, 1500, and 2500, a.u., smaller values allow for a more varying deformation field) to account for B_0_-inhomogeneity variation across subjects. The distortion corrected dataset with the highest mutual information (MI)^22^ between corrected positive and negative PE trajectory volumes was determined to be the most correct for each subject. The MI metric explains the amount of information one volume contains about the other.

The implementation of RPG used in this study differed from that originally described by Holland et al. in that input volumes (positive and negative PE trajectories) were resampled to a resolution of 2×2×2 mm^3^. In the authors’ experience, results are consistently better when voxel size is isotropic, rather than anisotropic. Displacement fields estimated from isotropic volumes were then properly scaled to the original voxel size and applied to the positive and negative PE direction volumes. The GE product RPG is similarly based on the original technique by Holland; however, the GE implementation is specific for neuroimaging applications and not validated outside the brain.

The RPG algorithm minimizes the difference between the corrected positive and negative PE trajectory volumes without any input from distortion free anatomical images. To evaluate the performance of the distortion correction algorithm, MI was estimated between T_2_ FSE images and positive PE volume before and after distortion correction. In addition, the average, median, mode and 95^th^ percentile absolute deformation between original and distortion corrected b=0 s/mm^2^ volumes were computed. Results were reported within the constrained FOV for both the full and reduced-FOV datasets.

Mutual-information values were compared using two-way repeated-measures analyses of variance (ANOVA) with Sidak post hoc tests to identify significant differences between EPI modality (full vs reduced-FOV) and the effect of RPG distortion correction (before vs after). The threshold for significance (α) was set at 0.05 for all analyses. Statistical analyses were performed using Prism software (version 7 for Mac OS X, GraphPad Software, La Jolla, CA, USA). In addition, two-tailed paired t-tests were used to evaluate whether differences existed between the performance of RPG on full and reduced-FOV EPI. Bonferroni correction was used to adjust p-values for multiple comparisons. All data are reported as mean ± standard deviation values.

## Results

Uncorrected and distortion corrected images and estimated displacement field for breast phantom data are shown in **Figure 1**. The initial and residual distortion was estimated using the grid of circles within the phantom on EPI and T_2_ FSE images (**Figure 2, Table 1**). Initial distortion was decreased by a factor of two on reduced-FOV EPI (0.27 ± 0.18) compared to full-FOV EPI (0.54 ± 0.46), after RPG correction the residual distortion was 0.35 times smaller in reduced-FOV (0.15 ± 0.07) than in full-FOV EPI (0.23±0.14). RPG improved distortions within the breast phantom structure for both full and reduced-FOV EPI. However, the performance of RPG was limited in full-FOV EPI near the edges of both sides of the breast phantom (asterisks in **Figure 1**). Average and 95^th^ percentile displacement post-distortion-correction for full-FOV EPI data were 0.2±0.3 cm and 0.7 cm, and 0.1±0.1 cm and 0.3 cm for reduced-FOV EPI data.

**Figure 1.**
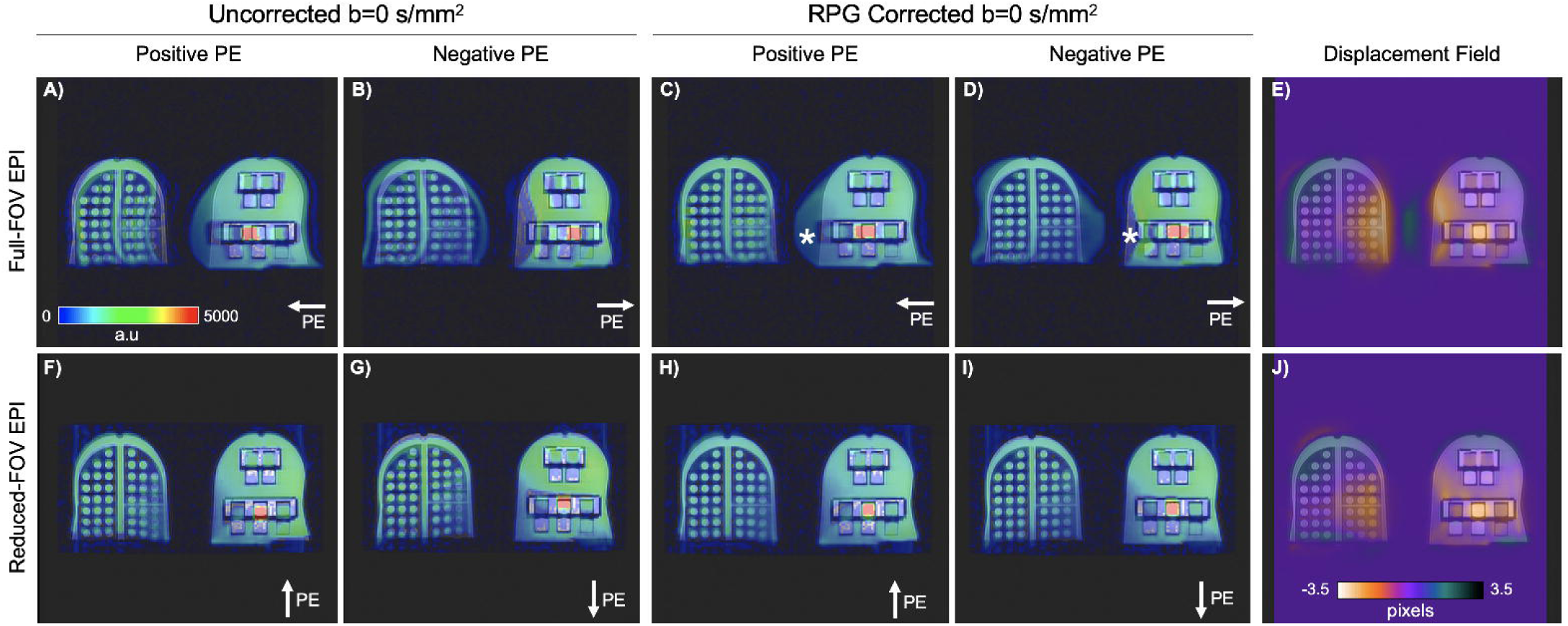
Overlay of positive and negative phase encoding (PE) trajectory polarities of both full (A, B) and reduced-FOV (F,G) EPI on T_2_-weighted fast spin echo (FSE) images of a breast phantom. The solution in the phantom interstitial space (shades of green) is fibroglandular tissue mimic consisting of 35% corn syrup in deionized water. High signal region (red in overlay) is 25% polyvinylpyrrolidone used to mimic tumor diffusion properties and the low signal regions (blue) are fat mimic (grape seed oil). Full (C,D) and reduced-EPI data (H,I) were distortion corrected using the reverse polarity gradient (RPG) method. Resulting displacement fields used to correct these data are shown (E,J). The RPG algorithm performs well when unwarping regions within the breast phantom structures as observed in the distortion plate (grid of circles spaced at constant intervals). However, distortion correction is limited on the medial region of the phantom (asterisks) in full-FOV EPI. a.u. = arbitrary units; PE = phase encoding.

**Figure 2.**
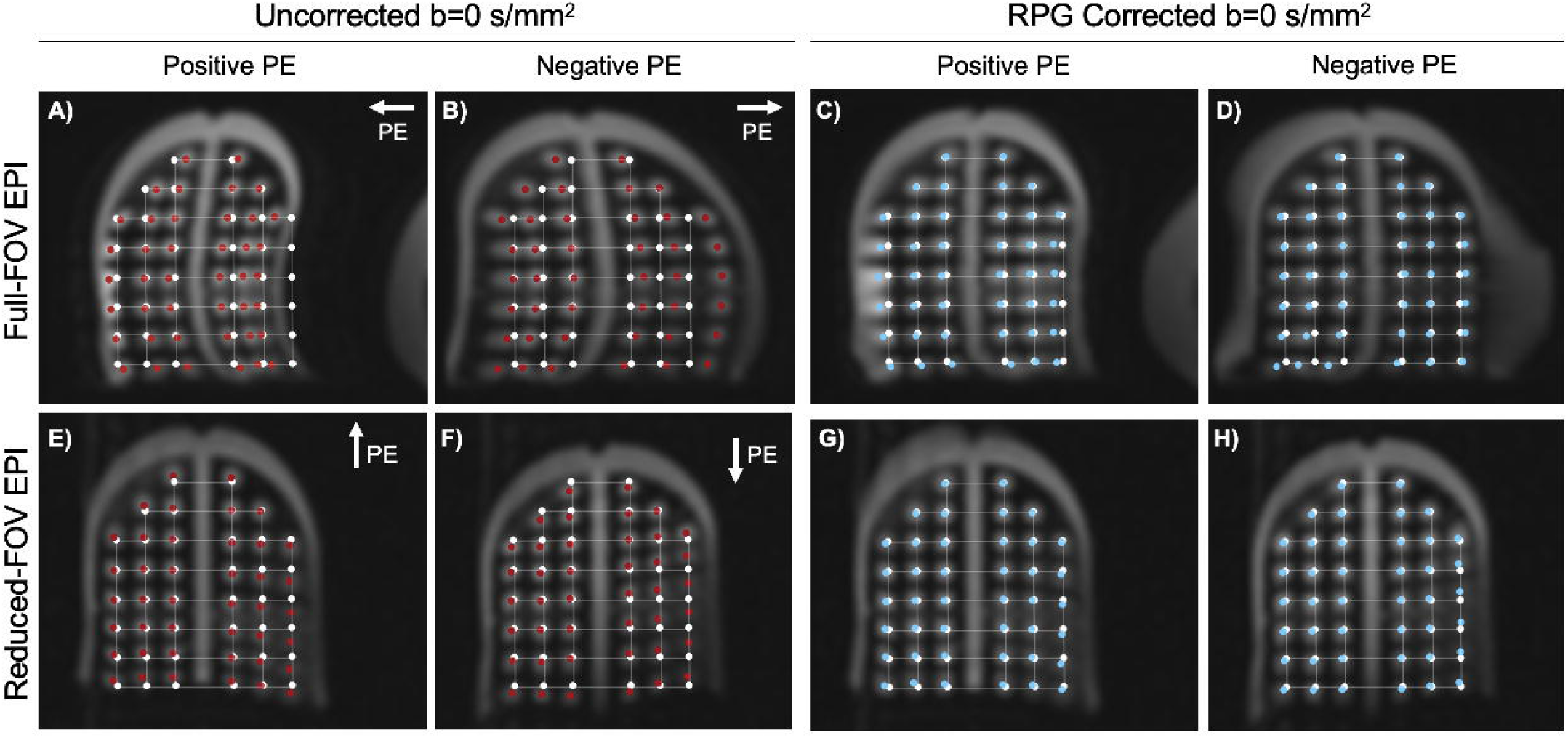
Full-(A-D) and reduced-FOV EPI (E-H) before (A, B, E and F) and after (C, D, G and H) distortion correction with the reverse polarity gradient (RPG) method of a breast phantom. White points indicate the location of the center of each circle on the polycarbonate grid on the T_2_-weighted fast spin echo (FSE) image. Red and blue points indicate the center (maximum signal intensity) of each circle on full and reduced-FOV EPI data before and after distortion correction, respectively. PE = phase encoding.

Representative *in vivo* cases are shown in **Figure 3** for women with different breast shapes. There was a significant (p<0.0001) increase in mutual information, MI, between EPI and T_2_ FSE images after RPG distortion correction of 13±8% and 8±6% for both full and reduced-FOV data, respectively. Overall, the MI between full-FOV EPI and T_2_ FSE images was higher (p<0.0001) than that between reduced-FOV EPI and T_2_ FSE images. This is attributed to higher signal-to-noise ratio (SNR) in full-FOV EPI data. The mode of the displacement field between uncorrected and corrected images were 0.2±0.1cm and 0.1±0.1cm for full and reduced-FOV EPI datasets, respectively. In contrast, the 95^th^ percentile was 3.0±1.0cm and 1.6±0.8cm for full and reduced-FOV EPI. This is in good agreement with the expected distortion reduction between both methods as the PE direction FOV reduction factor was 2.

**Figure 3.**
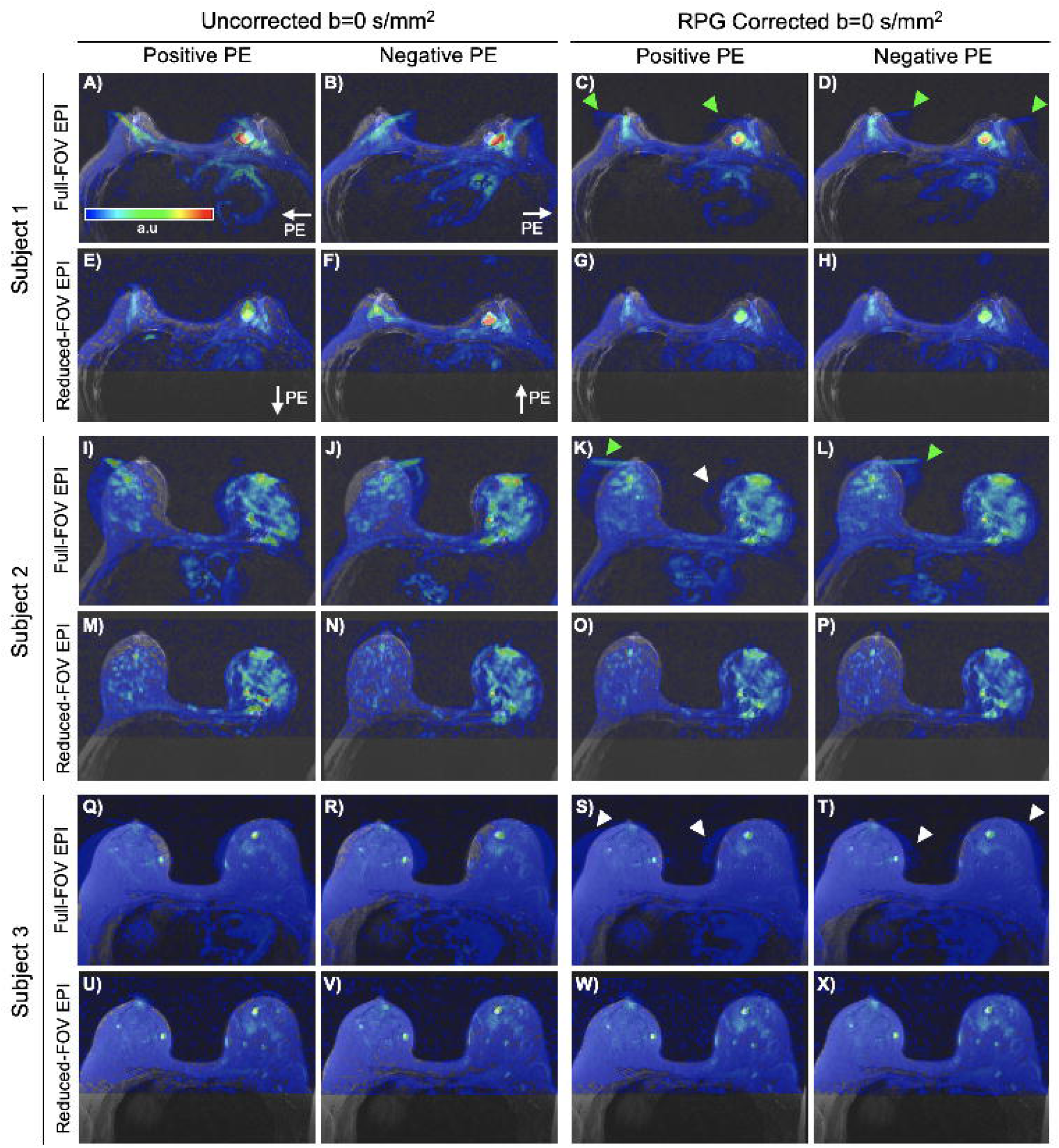
Overlay of uncorrected and RPG corrected full- and reduced-FOV EPI data on T_2_-weighted fast spin echo (FSE) images in three participants. Data were acquired in the positive (first column) and negative (second column) phase encoding (PE) trajectory polarities and were both corrected for geometric distortions using the reverse phase gradient (RPG) retrospective approach (third and fourth columns). Arrows indicate residual distortion in the breast fatty tissue (white) and nipple region (green) after distortion correction with RPG. a.u. = arbitrary units; PE = phase encoding.

Visual inspection of distortion corrected EPI data revealed that this version of RPG improves distortion correction when high-signal features (e.g. glandular tissue, tumor lesions) are present in b=0 s/mm^2^ images. However, despite improved agreement between EPI and T_2_ FSE images, RPG displayed limited capability to correct for geometric distortion in the fatty tissues in full-FOV EPI data, probably due to low signal intensities resulting from fat suppression, leading to the small mean and median distortion correction displacements achieved with RPG (**Table 2**). An important difference between corrected full and reduced-FOV EPI data was observed in the nipple area. In the majority of the cases, it was impossible for RPG to completely correct for the distortion artifact in the nipple region in the full-FOV EPI data (**Figure 3**, green arrows) and depending of breast shape the artifact was too severe in the anterior portion of breasts. This was not observed in reduced-FOV EPI data.

## Discussion

CE-MR is the most sensitive breast imaging modality and is important in the assessment of extent of disease and response to neoadjuvant chemotherapy. Cumulative exposure of intravenous contrast agents form depositions of such agents in brain with unknown long term sequalae.^6,7^ Diffusion-weighted imaging has shown potential to complement current surveillance and diagnostic standard of care for breast cancer.^23-25^ A critical limitation for the widespread use of DW-MRI in breast MRI is the distortion artifacts that greatly reduce its utility. Readily available and efficient prospective and retrospective strategies may be implemented to minimize and correct these artifacts. In the present study, we evaluated the performance of prospective distortion reduction methods compared to conventional EPI, and in combination with a custom version of a retrospective distortion correction method (RPG) for breast MRI. Overall, improved agreement was found between distortion corrected EPI of opposite PE trajectory polarities.

Other advanced MRI methods have been implemented to minimize distortion artifacts in breast EPI data. For instance, Taviani et al. incorporated parallel imaging and reduced-FOV DW-MRI to successfully produce high-resolution images (0.8×0.8×4.0mm^3^) with minimal distortion artifact.^13^ Similarly, Lee et al. developed a strategy in which B_0_-mapping is performed prospectively during pre-scan adding ∼2 minutes of scan-time. B_0_-inhomogeneities are then minimized by dynamically updating per-slice shimming through EPI acquisition, resulting in reduced distortion artifacts.^10,12^ The performance of several prospective and retrospective distortion correction strategies for breast DW-EPI were compared by Hancu et al.^12^ Similar to the results of the present study, findings by Hancu et al. suggest that a combination of prospective and retrospective strategies (in their case, acquisition with dynamic per-slice shimming^10^ and post-processing with RPG^19^) may be necessary to produce breast EPI data with minimal geometric distortion.^12^

The performance of RPG in correcting the geometric distortion from full-FOV DW-MRI breast data (acquired with parallel imaging) was previously evaluated by Teruel et al^11^. Results showed improved spatial agreement between diffusion-weighted and anatomical images.^11^ However, the work by Teruel et al. was performed with images collected on a single breast in the sagittal plane, which is not representative of typical clinical breast imaging. In contrast, data in the present study were acquired in the axial plane, which is the standard in clinical breast MRI protocols, offering a more translational evaluation of RPG performance.

In the present study, it was observed that RPG corrects distortion artifacts well when high-signal features are present in the positive and negative direction PE images. In other words, RPG requires features well above background noise level to properly register the positive and negative direction PE images (step 2 of the distortion correction algorithm in methods section). Thus, to produce DW-EPI data with the maximum clinical utility for patients with and without high intensity signal features in breasts, the use of reduced-FOV EPI and RPG together with parallel imaging may be of benefit. Parallel imaging reduces the magnitude of the distortion artifacts by decreasing the lines collected in PE direction (similar to reduced FOV acquisitions), thus reducing the readout duration.

The multi-shell DW-EPI pulse sequence used in the present study incorporates the collection of b=0 s/mm^2^ in both PE trajectory polarities, while non-zero b-value DW-MRI data were collected only in the positive PE trajectory polarity. The resulting RPG deformation field is then used to unwarp diffusion-weighted data to correct for geometric distortions before computing DW-MRI estimates. Full and reduced-FOV EPI data were collected with identical pulse sequence parameters, except for the PE direction. Hence, distortion artifacts were of different magnitude and prominent along different axes. A relevant and common example of this occurs in the nipple and medial regions of breasts, where the distortion artifact is, in most cases, uncorrectable with RPG on full-FOV EPI data. In contrast, the reduced-FOV EPI data, the nipple and medial regions distortions were decreased and then corrected with RPG. The reduced-FOV EPI methods produce images with decreased distortion artifact magnitude. An additional advantage of reduced-FOV EPI is that chemical shifts are decreased by the same factor as the distortion predominantly occur in the slice direction. However, reduced-FOV EPI images exhibit decreased SNR compared to conventional EPI, as only a fraction of k-space data is collected. Further studies evaluating the diagnostic value of reduced-FOV EPI remain to be performed.

A potential disadvantage of using reduced-FOV EPI in breast applications is limited axillary coverage. However, good visualization of the axilla is attained, as the excluded FOV of the chest for most patients is posterior to the axilla. In large patients, an alternative may be to reduce the PE direction FOV by a smaller factor; however, this may come at a cost of more modest improvement in the magnitude of the distortion artifacts. Unfortunately, as coverage in the PE direction (A/P) increases, images will potentially become more severely affected by cardiac and breathing motion artifacts. In such cases, the use of full-FOV EPI with PE in the L/R direction may be advantageous. In summary, the work presented here demonstrates that RPG decreases measurable distortions due to B_0_ inhomogeneity. Based on the present work, breast EPI data with reduced distortion artifact can be readily achieved by combining prospective and retrospective correction strategies such as reduced-FOV EPI and RPG.

## Data Availability

Data may be shared upon request.

## Acknowledgements

GE Healthcare, RSNA Research Resident Grant, and California Breast Cancer Research Program Early Career Award.

